# THE USE OF AN OBJECTIVE METHOD (CONTINUOUS EXOSOMATIC ELECTRODERMAL ACTIVITY WITHOUT EXTERNAL STIMULI) TO EVALUATE PATIENTS WITH HYPERHIDROSIS UNDERGOING VIDEO-ASSISTED SYMPATHECTOMY

**DOI:** 10.1101/2024.06.16.24309007

**Authors:** Rafael José Silveira, Carolina Carvalho Jansen Sorbello, Nelson Wolosker, José Ribas Milanez de Campos, João José de Deus Cardoso, Alexandre Sherlley Casimiro Onofre

## Abstract

2.

**Objective:** To objectively assess continuous exosomatic electrodermal activity without external stimuli in primary hyperhidrosis patients, before and after sympathectomy and compare it with responses to clinical investigation questionnaires.

**Method:** In a prospective study, 28 participants were divided into two groups. The first group consisted of 18 patients with palmoplantar hyperhidrosis who underwent sympathectomy on the fourth and fifth costal arches, bilaterally and sequentially. The second group, serving as a control, consisted of 10 patients. The assessment involved 2 self-explanatory questionnaires; the control group completed the questionnaires once, while the sympathectomy group completed them before surgery, one day and 30 days later. Sweating was objectively evaluated by measuring electrodermal activity (EDA) using the MP36R biosensor from Biopac Systems Inc. USA. This was done by carrying out measurements before surgery, on the first postoperative and on the thirtieth postoperative using an exosomatic technique and a constant electrical flow. Measurements were taken sequentially from the hands and feet, for 5 uninterrupted minutes at each site, after 10 minutes of rest, in a comfortable sitting position, without external stimuli, and in an air-conditioned environment. The study also collected anthropometric, clinical, and surgical data, and no significant sociodemographic differences were observed between the groups.

**Results:** In the group that underwent thoracic sympathectomy, there was a significant improvement in quality of life and a reduction in palmar and plantar sweating, as assessed by the questionnaires. Electrodermal activity showed significantly higher levels in the hands and feet of patients with hyperhidrosis compared to the control group during the preoperative assessment. After surgery, there was a reduction in electrodermal activity in the hands, and 100% of the sample analyzed showed a decrease in sweating. As for the evaluation of the feet, 67% of the patients reported a reduction in sweating, and 44% showed a statistically significant decline in EDA.

**Conclusion:** Continuous exosomatic electrodermal activity without external stimuli is a suitable method for assessing patients with palmoplantar hyperhidrosis, with appropriate clinical correlation compared to the questionnaire’s answers quantifying sweating and quality of life.

## 3. INTRODUCTION

Primary hyperhidrosis (HH) is characterized by excessive sweating unrelated to external triggers or body temperature regulation (1). It is often associated with dysfunction of the sympathetic nervous system (SNS) (2). This condition most commonly affects the palms of the hands, soles of the feet and axilla, but can also be present on the face, trunk, and other parts of the body. Increased sweating can lead to embarrassment and have a negative impact on a person’s social and psychological well-being (3), causing distress and reducing the quality of life (QoL) for those affected. Diagnosis is primarily based on clinical assessment, using questionnaires to measure sweating and evaluate its impact on the patient’s QoL. The only definitive treatment is surgery, however, symptoms can be managed with the use of medications such as oxybutynin (4,5).

Video-assisted thoracoscopic thoracic sympathectomy (VATS) is considered one of the best treatment options for localized forms of HH due to its effectiveness and safety (6). The level of intervention in the sympathetic chain is the main factor influencing these results (7).

It is essential to evaluate the degree of sweating and QoL when investigating the disease.(8) Patients are typically given simple, easy-to-understand questionnaires to assess the intensity of sweating in the main affected areas of the body using scales like the Hyperhidrosis Disease Severity Scale (HDSS) (9). These questionnaires also help to relate the volume of sweating to the repercussions on the interviewees’ routine activities (10). Questionnaires are cost-effective and non-invasive; however, they rely on the patient’s personal interpretation and cognitive abilities, which can introduce subjectivity.(11,12).

The objective methods for quantifying sweat already described in the literature are not commonly used in medical practice. Some of the best-known objective methods include transepidermal sweat dosage and pad glove (which quantifies sweat transferred to a specific type of glove) (13). However, the variability and lack of specificity of the measurements, complex techniques, short measurement times, and the cyclical nature of HH, make it challenging to use these measurements in clinical practice.

The EDA represents the electrical properties of the skin, which result from sympathetic activity and cause the release of sweat. This alters the concentration of salts in the cells and creates an electrical potential (14,15). This tool has been extensively used in psychological and behavioral studies as well as evaluating stress responses (16). There are 2 methods for measuring EDA: exosomatic (involving the application of an external electric current) and endosomatic (without an external electric current) (17). Both techniques have been used in the past to study patients with HH, yielding different results (18). However, the continuous measurement of EDA by exosomatic technique and without external stimulus (EDAcw) has not been used to study patients with HH undergoing sympathectomy.

In 2008, Tronstad et al. introduced a portable, non-invasive instrument capable of continuously measuring exosomatic EDA. This device was connected to a computer system and represented a significant advancement in medical research (19).

The aim of this study was to prospectively analyze the application of the exosomatic technique without external stimulus, with the use of a portable device to continuously measure EDA in individuals with palmoplantar HH before and after surgical treatment, and compare it with data from individuals without the disease. Additionally, the study correlated these findings with established clinical diagnostic methods such as HDSS and QoL.

## 4. CASUISTRY AND METHOD

From January 2023 to January 2024, a prospective study was conducted involving 28 participants. The study measured the intensity of sweating in 18 patients with palmoplantar HH who had undergone VATS, and 10 individuals without HH, using EDAcw. The study was approved by the Human Research Ethics Committee of the Federal University of Santa Catarina - UFSC (CAAE: 43287321.8.0000.0121), and all participants provided informed consent.

The participants were divided into 2 groups: the sympathectomy group, which comprised 18 (64.3%) patients with palmoplantar HH who had undergone bilateral sympathectomy, and the control group, which consisted of 10 (35.7%) individuals without HH and without surgical intervention. Both groups had similar demographic characteristics, with a majority of female patients (72.2% versus 80.0%), who were in their third decade of life (25 versus 22.5 years) and with a body mass index of less than 25 (23.1 versus 22.1Kg/m2).

Patients in the sympathectomy group underwent bilateral sympathectomy (right side followed by the left side) on the fourth and fifth costal arches (R4/R5). The procedure involved using monopolar electrocautery without a direct approach to the thoracic ganglion. The surgery was performed under general anesthesia, with orotracheal intubation using a double-lumen tube, and non-invasive cardiovascular monitoring was conducted. Incisions of 0.5 - 1.0 cm were made in the fifth intercostal space, in the anterior axillary line to pass the optical system, and in the third intercostal space, in the middle axillary line, to introduce the electrocautery after local infiltration with Ropivacaine. The procedures were carried out by the same team throughout the study period, using a standardized surgical technique.

All patients except one were discharged on the first day after surgery. The remaining patient was discharged on the second postoperative day due to a residual pneumothorax and the need for pleural drainage. Compensatory hyperhidrosis (CH) was mild to moderate in ten patients (55.6%), severe in one patient (5.5%), and absent in seven patients (38.9%).

The participants underwent a clinical assessment, completed questionnaires, and had their EDAcw measured on different occasions. The control group underwent these assessments once, while the sympathectomy group underwent them on three occasions: before surgery, one day after, and 30 days after the operation. The same investigator conducted all assessments.

To measure the intensity of palmar and plantar HH, we used the Hyperhidrosis Disease Severity Scale (HDSS), which is graded numerically from 1 to 4, representing the lowest to the highest intensity of sweating.

To assess QoL, we used the quality of life questionnaire (20). The patients completed the QoL assessments independently, without any influence from the doctor. The preoperative QoL was categorized into five satisfaction levels based on the total points obtained from the questionnaire: very poor (total score > 84), poor (total score 69 to 84), good (total score 52 to 68), very good (total score 36 to 51), and excellent (total score 20 to 35). After the surgery, the QoL assessment was repeated thirty days post-operatively. The QoL assessment was repeated and the patients were also classified into one of five different levels of satisfaction, according to their score: much worse (total score)> 84; slightly worse (total score 69 to 84); unchanged (total score 52 to 68); slightly better (total score 36 to 51); and much better (total score 20 to 35).

For the assessment of EDAcw, we used the MP36R biosensor from Biopac Systems Inc. USA before and after the surgery. To measure EDAcw we used the exosomatic technique with a constant electrical flow of 0.5 Voltz. Disposable electrodes were placed on the thenar and hypothenar regions of the hands and the middle third of the medial surface of the feet. Measurements were taken after 10 minutes of rest, in a comfortable seated position, on the right and left hands followed by the right and left feet, sequentially, for 5 uninterrupted minutes in each location, without any specific stimulus, in a quiet environment with a temperature of between 21oC to 23oC and air humidity of 60% to 65%. The electrophysiological signals captured were digitized and transmitted to a computer system via a USB cable for analysis using the AcqKnowledge software. The control group underwent electrodermal measurements only once.

EDA was estimated in two ways: through mean skin conductance (MSC) and skin conductance area (SCA) in the hands and feet at three different times: before surgery, on the first postoperative day, and on the thirtieth postoperative day.

First, we analyzed the responses to the QoL and HDSS questionnaires and measured the average EDAcw and area in the control group and the sympathectomy group during each period analyzed (pre-surgery, first day after surgery, and thirtieth day after surgery). Then, we individually studied EDAcw in the subgroups of patients with improved sweating on the hands, followed by the group with significant improvement in sweating on the feet, the group with a slight improvement in sweating on the feet, the group with maintained sweating on the feet and finally the group with worsening sweating on the feet after surgery.

### Statistical analysis of data

We first conducted a statistical analysis on the clinical data, followed by an analysis of the responses to the questionnaires (HDSS and QoL), and, finally, an examination of the EDA data.

We used the The Shapiro-Wilk test to check for normality of the data. Numerical variables were described using measures of central tendency and dispersion, while categorical variables were described using absolute and relative frequencies. The Student’s t-test was used to analyze independent samples, and the Mann-Whitney test to analyze the difference between groups. The Wilcoxon and Friedman tests were employed to compare two and three different moments, respectively. We considered p-value of ≤ 0.05 to indicate statistical significance. All data was analyzed using the R programming language version 4.2.1

## 5. RESULTS

The answers to the questionnaires and the EDA measurements are shown in Table 1.

**Table 1:** The answers to the questionnaires (QoL and HDSS) and measurement of the EDA in the control group and the sympathectomy group in each period analyzed (preoperative, first day after surgery and thirtieth day after surgery).

| Group | Control | Sympathectomy |  |  |
| --- | --- | --- | --- | --- |
| Variables |  | Before Surgery | 1 Day After Surgery | 30 Days After Surgery |
| **Quality of life** | - | 87,5 (80,8 – 93,3) | - | 22,5 (20,8 – 28,5) |
| **HDSS** |  |  |  |  |
| **Hands** | 1 | 4,0 (3,0 – 4,0) | - | 1,0 (1,0 – 1,0) |
| **Feet** | 1 | 3,0 (3,0 – 4,0) | - | 2,0 (1,0 – 2,3) |
| **EDA (average)** |  |  |  |  |
| **Hands** | 5,15 (2,0-2,8) | 11,3 (6,6 – 13,3) | 0,3 (0,2 – 0,5) | 1,3 (0,4 – 3,9) |
| **Feet** | 2,35 (1,6-1,9) | 11,2 (5,9-12,4) | 0,3 (0,2-0,5) | 1,2 (0,4-3,7) |
| **EDA (area)** |  |  |  |  |
| **Hands** | 186,3 (53,2 – 390,7) | 388,4 (284,3 – 630,2) | 5,6 (1,1 – 17,9) | 96,1 (20,0 – 130,0) |
| **Feet** | 24,95 (10,8 – 156) | 352,1 (278,7-647,7) | 6,2 (1,3-18,4) | 92,9 (18,0-128,6) |

| Group | Control | Sympathectomy |  |  |
| --- | --- | --- | --- | --- |
| Variables |  | Before Surgery | 1 Day After Surgery | 30 Days After Surgery |
| Quality of life | - | 87,5 (80,8 – 93,3) | - | 22,5 (20,8 – 28,5) |
| HDSS |  |  |  |  |
| Hands | 1 | 4,0 (3,0 – 4,0) | - | 1,0 (1,0 – 1,0) |
| Feet | 1 | 3,0 (3,0 – 4,0) | - | 2,0 (1,0 – 2,3) |
| EDA (average) |  |  |  |  |
| Hands | 5,15 (2,0-2,8) | 11,3 (6,6 – 13,3) | 0,3 (0,2 – 0,5) | 1,3 (0,4 – 3,9) |
| Feet | 2,35 (1,6-1,9) | 11,2 (5,9-12,4) | 0,3 (0,2-0,5) | 1,2 (0,4-3,7) |
| EDA (area) |  |  |  |  |
| Hands | 186,3 (53,2 – 390,7) | 388,4 (284,3 – 630,2) | 5,6 (1,1 – 17,9) | 96,1 (20,0 – 130,0) |
| Feet | 24,95 (10,8 – 156) | 352,1 (278,7-647,7) | 6,2 (1,3-18,4) | 92,9 (18,0-128,6) |

a Values expressed as median and interquartile range (P25-75); b Wilcoxon test; c values expressed as mean and standard deviation; d paired t-test.
HDSS = Hyperhidrosis Disease Severity Scale

**- Clinical assessment:**

We observed that the intensity of sweating on the hands, as measured by the HDSS before surgery, was highest in the sympathectomy group (4.0) and decreased to a minimum level postoperatively, a value similar to that of the control group [z = -3.804; p < 0.001].

For the feet, the average HDSS was 3.0 preoperatively and improved to 2.0 [z = -3.007; p < 0.01] after surgery, even surpassing that of the control group.

Furthermore, there was an improvement in the patient’s QoL after surgery. Patients reported very poor QoL before surgery, and after sympathectomy, they showed significant improvement [z = -3.726; p < 0.001].

**- Electrodermal activity analysis (EDA)**

### EDA in the hands

The average intensity of EDAcw in the hands of the 18 patients who had surgery is shown in Table 1 and illustrated in Figure 1. The study found statistically significant differences in EDAcw levels at three observation times: on the preoperative period, on the first day after the surgery, and on the thirtieth day after the sympathectomy. The comparative evaluation revealed differences between each moment; there was a difference between the preoperative period and the 1st PO day (p < 0.001), between the preoperative period and the 30th PO day (p = 0.02), and between the 1st PO day and the 30th PO day (p < 0.01).

**Figure 1:**
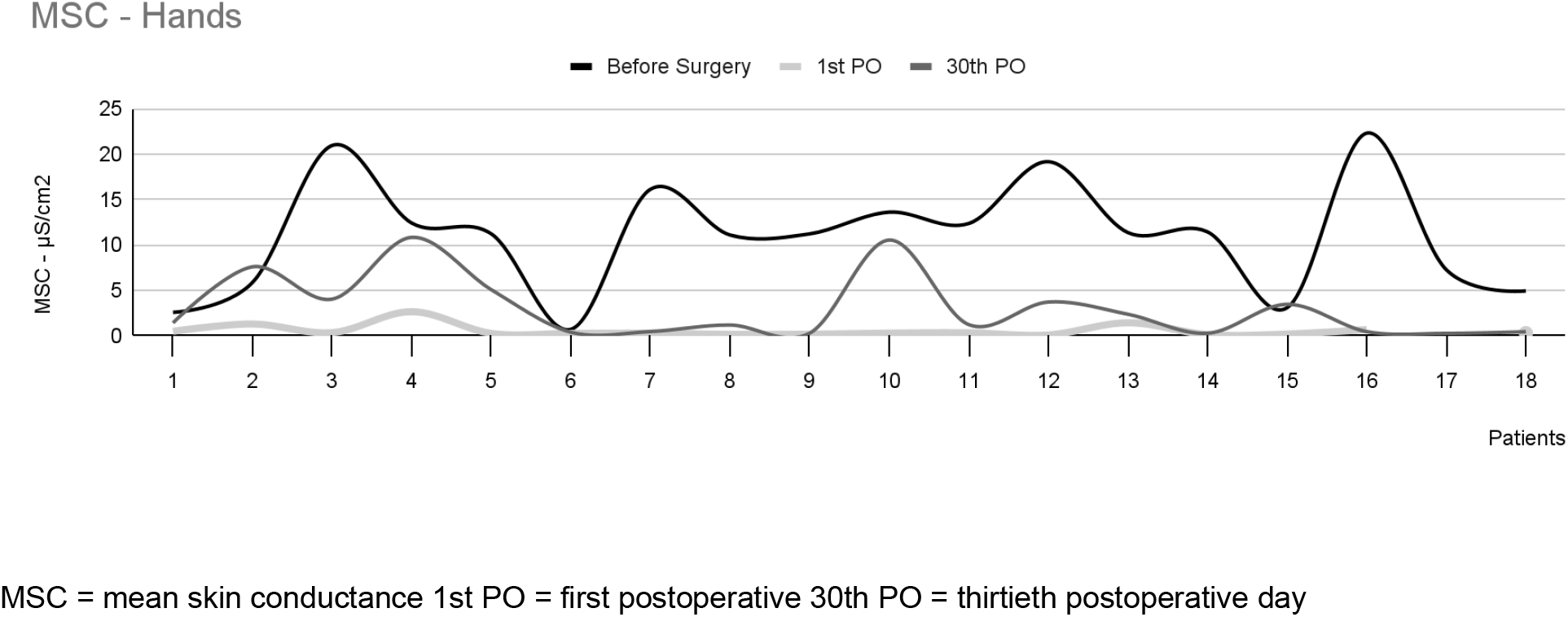
mean skin conductance in the hands of 18 patients with palmar hyperhidrosis.

The intensity of EDAcw in the hands, as measured by area, is described in Table 1 and illustrated in Figure 2. A statistically significant difference was observed among the three observation times [χ2(2) 28.444; p < 0.001]. When comparing each time point, differences were found between the preoperative period and the 1st PO day (p < 0.001), between the preoperative period and the 30th PO day (p = 0.01), and between the 1st PO day and the 30th PO day (p = 0.01).

**Figure 2:**
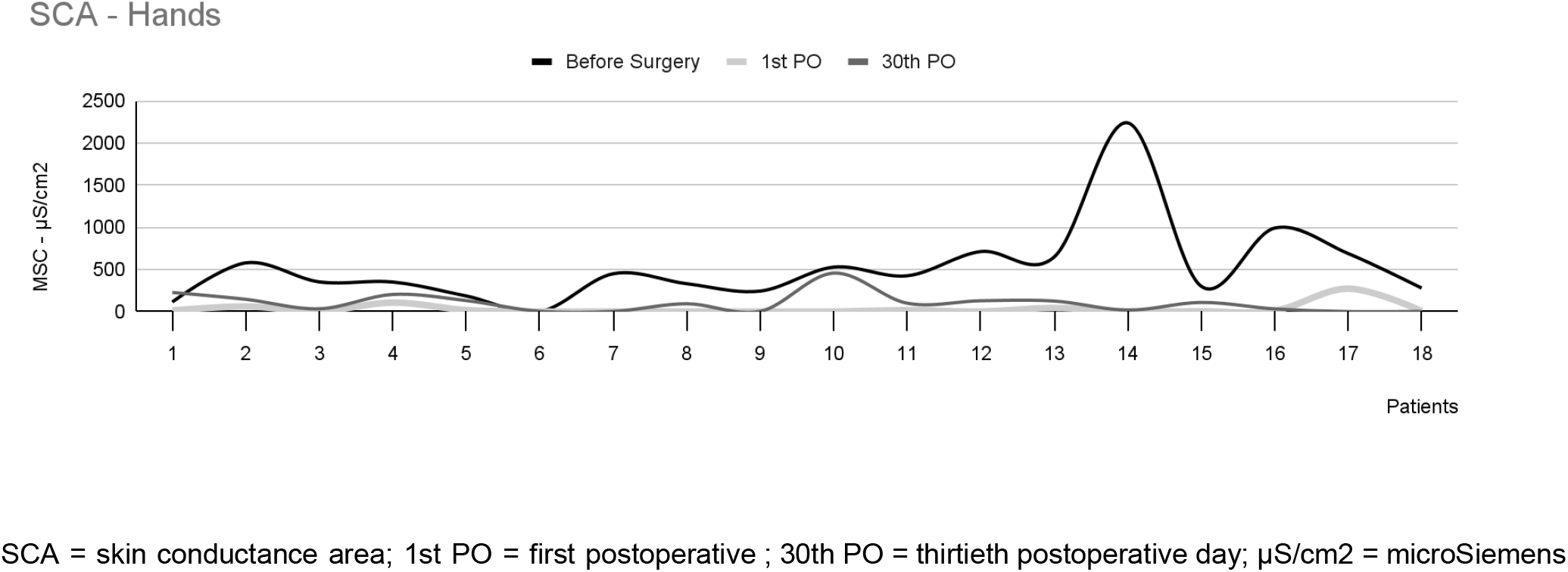
skin conductance area in the hands of 18 patients with palmar hyperhidrosis.

In the sample analyzed, only one patient (patient 12) experienced a slight decrease in sweating and showed a much smaller reduction in MSC between the preoperative period (13.6uS) and the 30th postoperative day (10.5uS), i.e. a 23% reduction in the intensity of EDAcw one month after surgery. The other 17 patients showed a reduction of approximately 97% in MSC over the same period. When we examined patient 12’s sweating using SCA, we found no significant difference between the measurements taken before the surgery (527.7uS/cm2) and 30 days after (456.7uS/cm2). However, there was a noticeable decrease in EDA in the 1st PO, indicating that the surgical procedure was performed correctly.

### EDA in the feet

The average intensity of EDAcw in the feet of the 18 patients who had surgery showed a statistically significant difference between two of the three observation times [χ2(2) 10.941; p < 0.01], which were between the preoperative period and the 1st PO day (p < 0.01) and between the preoperative period and the 30th PO day (p = 0.02). However, no significant difference was noted between the 1st PO and the 30th PO (p = 0.61).

The patients reported divergent results in terms of reduced plantar sweating after sympathectomy. Out of 18 participants who underwent surgery, 8 reported significantly reduced sweating. In these patients, the mean intensity of the EDAcw showed a statistically significant difference between the preoperative period and the 1st PO day (p = 0.05) and between the preoperative period and the 30th PO day (p = 0.02). However, there was no difference between the 1st PO and the 30th PO (p = 0.60), as shown in Figure 3. There was no statistically significant difference in the area of the EDAcw in this group.

**Figure 3:**
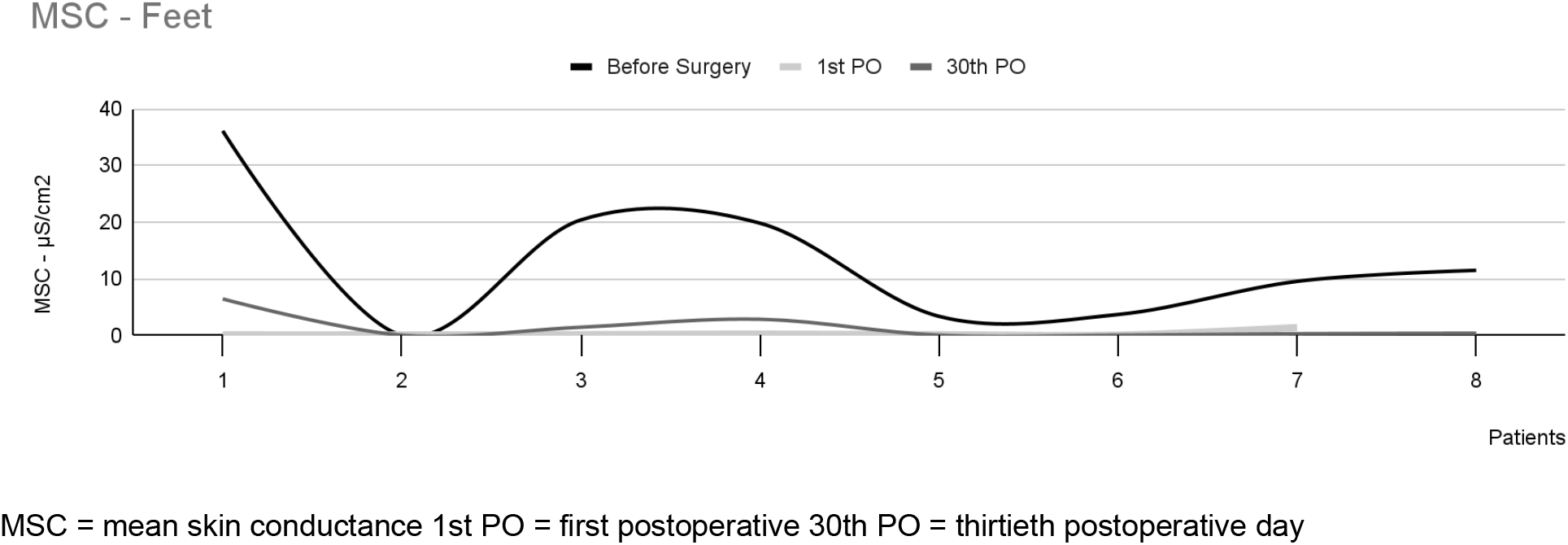
mean skin conductance in the feet of 8 patients with plantar hyperhidrosis and significant decrease in sweating.

**Figure 4:**
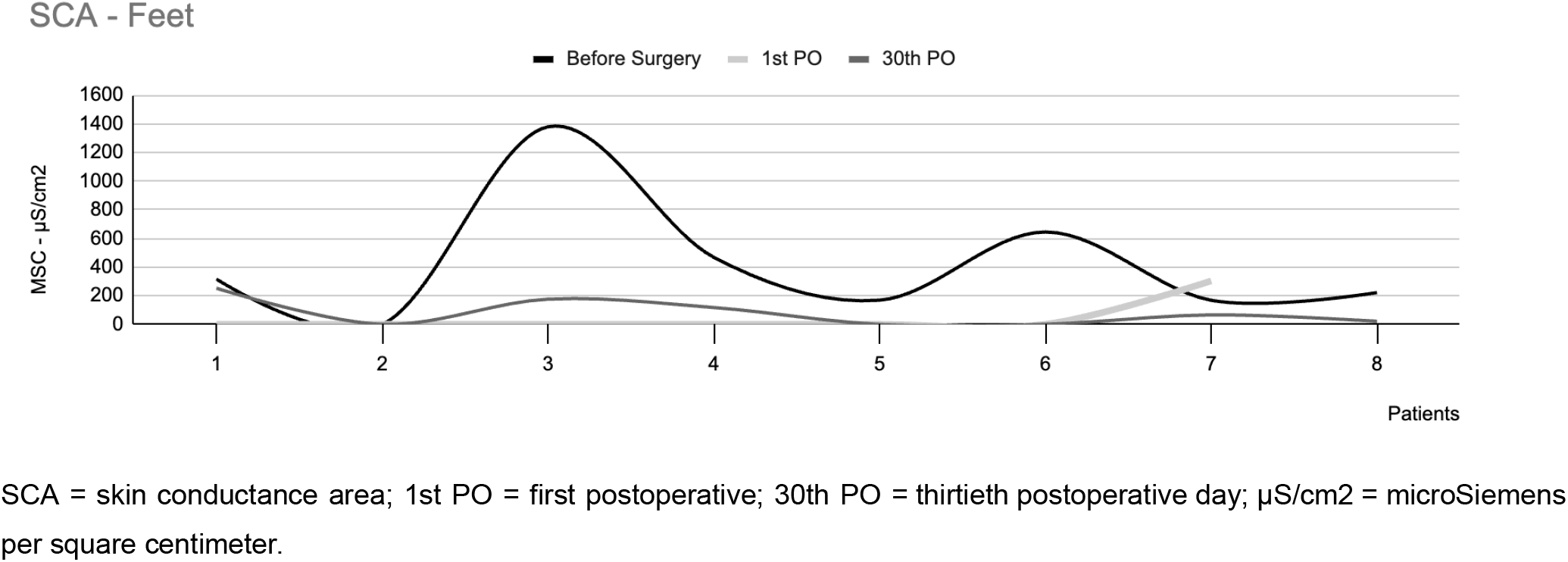
skin conductance area in the feet of 8 patients with plantar hyperhidrosis and significant decrease in sweating.

Of the remaining 10 patients, 4 reported a slight reduction in sweating. The mean EDAcw intensity and the EDAcw per area in this group showed no statistically significant difference between the three observation times [χ2(2) 2.000; p = 0.37], as shown in Figure 5.

**Figure 5:**
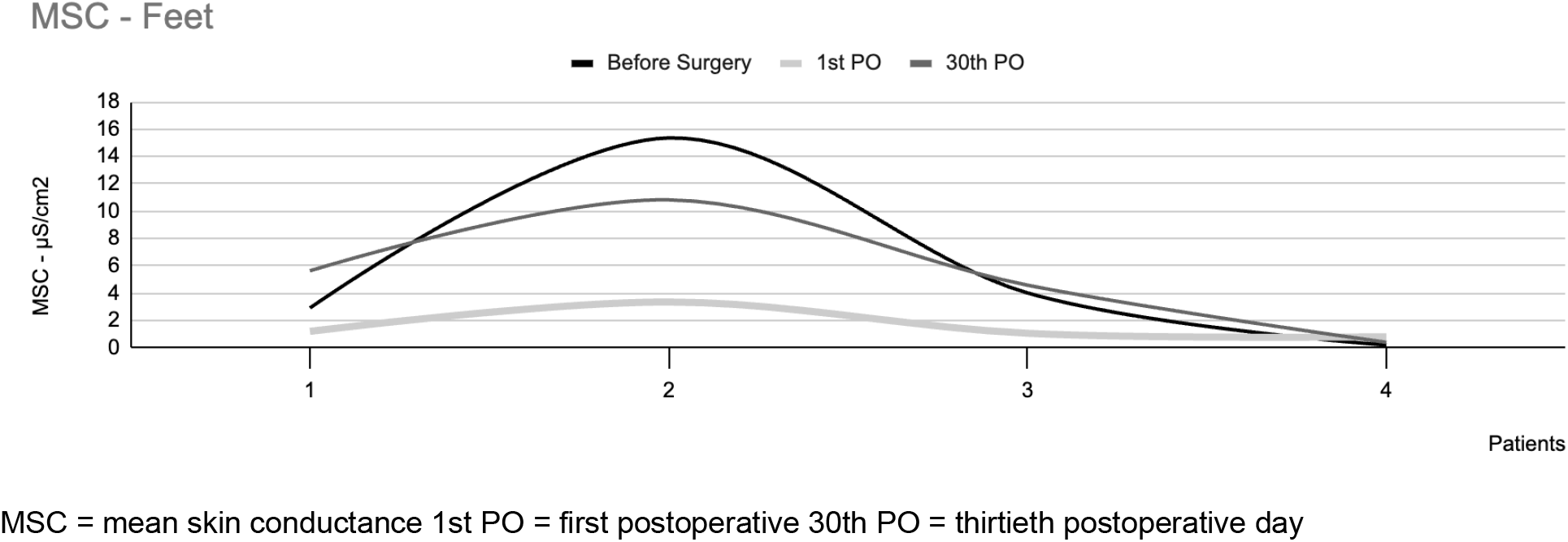
mean skin conductance in the feet of 4 patients with plantar hyperhidrosis and slight decrease in sweating.

For the 5 patients who did not show any clinical change in plantar sweating, the intensity of EDAcw measured by MSC showed no statistically significant difference between the three observation times [χ2(2) 5.200; p = 0.07], as shown in Figure 6.

**Figure 6:**
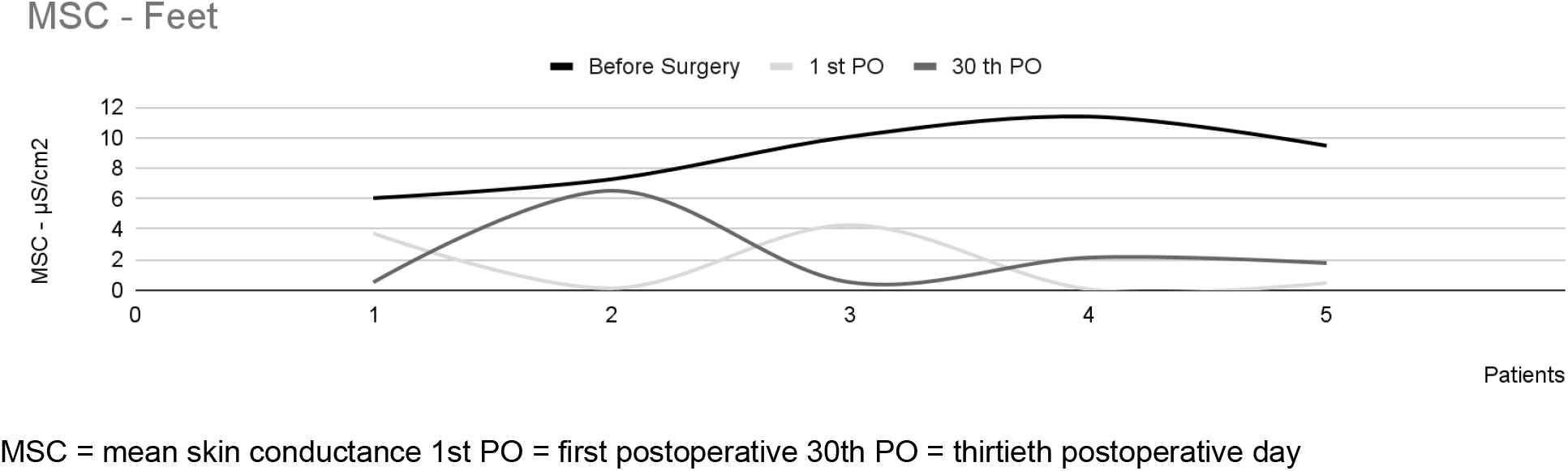
mean skin conductance in the feet of 5 patients with plantar hyperhidrosis and no change in sweating.

Only one patient reported an increase in post-operative plantar sweating and there was no clinical correlation with EDAcw measurement, as its measurements decreased in the post-operative period.

## 6. DISCUSSION

HH is a disease with a significant psychosocial impact on patients’ lives, directly affecting their QoL and mental health. Patients who undergo sympathectomy have shown considerable improvement in the QoL (21). The results of this study confirm the correlation between sympathectomy and a significant enhancement in patients’ QoL post-surgery.(22)

Decreasing in sweating is observed in approximately 70% of patients undergoing medical treatment (23,24), however, surgical sympathectomy is the only definitive treatment for the disease, resulting in an improvement in 90% of cases of palmoplantar HH (25). Since the 1990s, VATS has emerged as one of the best therapeutic options for localized HH (26). In our study, 97% of patients showed significant improvements, with only 1 patient not achieving satisfactory results.

The enhancement in patient’s QoL is the most important outcome of surgical treatment. It is directly related to the control of sweating and the intensity of compensatory hyperhidrosis (CH)(27), with the level of intervention in the sympathetic chain being the most relevant technical aspect for a satisfactory result .

The present study assessed VATS at the level of the 4th and 5th costal arches, and the post-operative analysis showed a significant reduction in palmoplantar sweating in practically all of the sample studied. Nicolini et al. , in a review article, pointed to a trend towards intervention at this level, mainly for treating palmar HH, which has been reaffirmed in several other studies (28).

In the context of plantar HH, VATS has shown inconsistent results. In a retrospective multicenter study, Chen et al. reported a 29.3% improvement in plantar sweating with thoracic sympathectomy (29), involving different surgical techniques. In another study, 40 patients underwent sympathectomy to treat HH. Approximately 45% of the patients had an decrease in plantar sweating, 32% maintained stable sweating, and around 22% experienced a worsening condition (30). This present study indicated an overall improvement in symptoms in roughly 66% of the patients. While VATS is not specifically targeted at plantar sweating, a significant percentage of patients may experience improvement.

The objective assessment of sweating was carried out using the EDAcw following the guidelines of the Society for Psychophysiological Research (31) to prevent possible extrinsic and intrinsic factors that could bias the results. The levels of EDAcw were significantly higher in the hands and feet in the group with HH compared to the group without HH, showing a strong clinical correlation. In a study by Manca et al. (32), EDA was analyzed using the endosomatic technique in 10 participants with HH and 10 without HH, aged between 26 and 50. They found that the group with HH exhibited more sympathetic responses and a shorter response latency time after sensory stimulation.

EDA is an objective measure of sympathetic activity in the eccrine sweat glands. Its electrophysiological origin is in the thermoregulatory center of the hypothalamus, but it is influenced by the entire cerebral cortex, especially the limbic system (33). Dooren et al. (34) evaluated the EDA of 17 participants in 16 dermatomes simultaneously for 3 minutes at each site, showing the relationship between this measurement and the density of the sweat glands and their level of stimulation. In a comparative study, Machado-Moreira et al. (35) found that it increased earlier than sweat levels, in 4 dermatomes of 14 participants, in response to an increase in body temperature, suggesting that this measurement precedes the presence of sweat.

Skin conductance (SC) is the key electrophysiological measurement analyzed in EDA. It can be obtained using exosomatic or endosomatic techniques, depending on whether or not an electrical current needs to be applied. This measurement is classified as skin conductance level, which represents slow and gradual variations in SC and is considered the baseline or “background” level of sympathetic activity, and skin conductance response or skin sympathetic response, which involves a sudden increase in the amplitude of skin conductance that can occur spontaneously (non-specific) or due to a specific stimulus (specific) (36).

Regarding the effects of thoracic sympathectomy on EDA, Lefaucheur et al. (37) showed a decrease in the amplitude of sympathetic responses in the hands on the side submitted to sympathectomy, as well as on the non-operated side. This suggests a neural plastic change at the central level of synaptic connections after surgery. However, there were no significant changes in the feet. On the other hand, Lewis et al. (38) did not find significant differences in EDA after VATS in 26 patients with HH.

In the present study, using the EDAcw, we detected a significant reduction in SC measurements in the hands and feet. This reduction was more pronounced on the first postoperative day compared to the thirtieth day, suggesting a neural plastic modification throughout the sympathetic chain after thoracic sympathectomy that changes over time. EDAcw measurements have proven to be effective in assessing sympathetic activity in the sweat glands and, consequently, the intensity of sweating, in a sensitive and accurate manner. Therefore, EDAcw can be considered a valuable tool for objectively evaluating patients with HH. The new instruments capable of measuring EDAcw in a continuous and simple way in various clinical scenarios and interfacing with a computer system make it possible not only to assess it in real time but also to store the data in a cloud. This enables review of the obtained measurements and analysis of other electrophysiological measurements. These features can contribute to a better understanding of the pathophysiology of HH and facilitate the classification and determination of the therapeutic approach, especially in relation to the different thoracic sympathectomy techniques.

## 7. CONCLUSION

Continuous electrodermal activity without external stimulus (EDAcw) proved to be a sensitive and efficient objective measure for evaluating patients with primary HH who underwent VATS in G4.

## Data Availability

All data produced in the present study are available upon reasonable request to the authors

## Notes

### Competing Interest Statement

The authors have declared no competing interest.

### Funding Statement

This study did not receive any funding

### Author Declarations

The study was approved by the Human Research Ethics Committee of the Federal University of Santa Catarina - UFSC (CAAE: 43287321.8.0000.0121), and all participants provided informed consent.

